# Understanding anaemia and stunting among young women in a rural setting of Indonesia

**DOI:** 10.1101/2022.12.14.22282506

**Authors:** Giyawati Yulilania Okinarum, Hardiningsih, Fresthy Astrika Yunita, Afroh Fauziah, Muhammad Hardhantyo

## Abstract

**Background:** Indonesian Ministry of Health stated that around 80% of Indonesian women aged between 15-24 years are anaemic, and 30% of children are stunted. The leading cause of this problem is poor quality food, including poor micronutrient quality, low dietary diversity and low intake of animal-source foods. Preconceptional young women who are anaemic and stunted have a risk of giving birth to stunted children later in life if their nutrition is not immediately improved. This study aims to have a deeper understanding of anaemia and stunting conditions among young women in the rural setting of Indonesia.

**Methods:** Twenty young women with the anaemic and stunting condition aged between 15 - 19 years were recruited through the randomly selected patient in the outpatient department of Public Health Center (Puskesmas) in the rural area of Gunungkidul, Yogyakarta from October to November 2022. Focus Group Discussion (FGD) was conducted with the following area early marriage, cause and effect of anaemia and stunting, and women health-seeking behavior. Interviews were audio-recorded, transcribed, translated, and analyzed thematically.

**Results:** Young women with anaemic and stunting condition are having powerlessness where they are unable in determining daily diet, arise from a low-income family and have no choice in their life course. They are suffering from financial difficulties and food insecurity, resulting in inadequate nutrition and stunting. They also believe that early marriage is a viable option for their own and their children’s well-being.

**Conclusion:** This study is the evidence of young women powerlessness in the rural area of Indonesia. The understanding could contribute to develop evidence-based, effective, and efficient policies and regulations. The existing health system needs to reinforce the support for young women to reduce risks in early life and improve their health across the life course. The intervention could include promoting access to nutritious foods, good hygiene, family planning education, and access to health facilities and services.

## Introduction

The global public health sector, including Indonesia’s healthcare system, is burdened by nutrition-related anaemia and stunting (1). This affect 1.62 billion people worldwide, most of them are children, adolescents, and women (2). According to the Ministry of Health, more than 80% of Indonesian women aged 15-24 years are anemic, and 30% of children are stunted (3). Another study in Indonesia included 645 elementary school students, it reported that 27% of them are anemic and 20% have stunted growth (4).

Anaemia is recurrently associated with malnutrition, which is characterized by reduced red blood cells. It is frequently accompanied by low Hb concentrations in the blood (5). A Kuwaiti study showed that children with stunted growth had a 2.3 times higher risk of anaemia than those without stunted growth (6). According to the other studies, anaemia and stunting affect all countries and have a negative impact on health as well as socioeconomic development (7,8), since nutrition is particularly important to adolescent nutrition on current and future adolescent health, future labor productivity, and the increase in the generation that would be brought into the world (9–11).

Malnutrition is listed as one of the consequences of food insecurity. According to a meta-analysis study, there is a general significant relationship between food insecurity and anaemia, the risk of iron deficiency anaemia, and stunted growth (8). Malnutrition among women has persisted over the last few decades, owing to the gender dimensions of food insecurity and the impact of their socioeconomic vulnerability on their disempowered personal nutrition (12). Household food insecurity is a significant correlate of anaemia in adolescent girls, hence the gender aspect of food insecurity and nutrition should be considered (13). For a considerable length of time, many developing countries have recognized the existence of gender disparities in income and food insecurity (14). Poverty and income inequality have a direct impact on the access and availability (15), its impact suffering in anaemia and stunting.

## Materials and methods

### Study design

An exploratory qualitative design with random sampling technique to recruit young Indonesian women in the rural area of Indonesia. This design has the advantage of drawing on the strengths of established qualitative methodologies and methods, allowing for the flexible application of techniques (16). Therefore, it is appropriate for investigating behavior, perspectives, feelings, and experiences. It essentially seeks to comprehend a phenomenon, a process, or the people involved’s perspectives and worldviews (17,18). An exploratory qualitative study was considered appropriate since the study aimed to have in-depth understanding of anaemia and stunting condition as seen through the eyes of young women living in rural area of Indonesia.

### Eligibility criteria for study participants

A total of twenty young women aged between 15 - 19 years were recruited through the randomly selected patient in the outpatient department of Public Health Center (Puskesmas) in the rural area of Gunungkidul, Yogyakarta from October to November 2021. Participants or participant’s parents gave consent for the interview and volunteered their time to have the Focus Group Discussion (FGD) with other participants. Additionally, the researchers were the only ones who knew the participant IDs, and database were kept all of the documentation, including signed consent forms, in a cabinet that was locked and participants may withdraw at any time without prejudice. Researchers H and AF measured the participants BMI and Hemoglobin (Hb) level. BMI measurements were performed with the portable scale and stature meter, and calculated by dividing body weight (in kilograms) with the square of height (in meter squares). BMI classification based on WHO criteria, thinnest was categorized for participant with < - 2SD and severe thinness for BMI < - 3SD, overweight for BMI > +1SD and Obesity for BMI >+2SD (19,20). Hemoglobin level were measured using a digital haemoglobinometer and further classified the anemic status, with the categorized: mild 11.0-11.9 gr/dL, moderate 8.0-10.9 gr/dL, and severe anaemia <8.0 gr/dL (21). Indonesia mandates a 12-year learning program; therefore, participants are categorized as quit school early if they do not complete their education for at least 12 years. “At school” category if participants are currently enrolled in education and graduated if they have completed high school. WHO height-for-age Z-score (HAZ) was used to assess nutritional status stunted was categorized for HAZ score < -2 SD, severely stunted for < - 3 SD and not stunted for others (20).

### Interview guide

Interview guidelines were developed to focus on the following area: early marriage, the cause and effect of anaemia and stunting, and the health-seeking behavior. Prior to the data collection, interview guidelines were face validated by the experts. Each item was reviewed, and several colleagues further requested to provide feedback on the questionnaire in order to ensure participants understood and accurately interpret the interview questions.

### Data collection

The FGD was conducted and further analyzed by the first author (GYO). Focus group discussions are held at the local village hall during working hours. Due to the widespread of COVID-19 pandemic in the area, the health protocol required to divided it into four FGDs, each of which consisted of five to six participants and enduring about two hours. The lead author audio-recorded all interviews and transcribed them verbatim (identifying features removed) in Indonesian. An independent assessor confirmed the audio recording and transcription (a confidentiality agreement). For data analysis, all transcripts were translated into English. A bilingual assessor independently assessed eight transcripts to ensure the accuracy of the translation, i.e., the narrative meaning, to evaluate the reliability of the translation into English and the back translation into Indonesian. In addition, independent assessors clarified perspectives of statements and phrases that could not be directly translated between languages.

### Analysis

Data were analyzed in six stages: familiarization with the data, generation of coded data, finding for themes, assessing themes, determining and emerging themes, and report production (22). NVivo 12 assisted in data management and the development of emergent iterative themes based on the semantic content of the codes. A reflexive approach enabled GYO to consider and discuss potential personal influence from her own biases throughout the study and analytical process (23).

To determine the study’s credibility, the researchers applied Lincoln and Guba’s four criteria (24). Triangulation and member checking were used to establish credibility, while the thick description was used to achieve transferability. To ensure dependability, field notes were taken throughout the study, and the advisory team served as auditors. The analysis’s veracity was established through an audit trail and method triangulation, which included both individual in-depth interviews and focus groups.

### Ethical considerations

This study was approved by the Research Ethics Committee of Faculty Medicine Universitas Sebelas Maret (Approval number: 88/UN27.06.11/KEP/EC/2021). The data collection procedures were meticulously designed to ensure confidentiality. All potential participants were informed verbally and in writing about the study’s methods, potential risks and benefits, and the duration of the study. All potential participants were also informed that their participation was entirely voluntary and that they could refuse or withdraw from the study at any time without consequence. When the students and their parents agreed, they were asked to sign a written informed consent form. Throughout the study, participants’ confidentiality and anonymity were maintained.

## Results

### Characteristics of study participants

All participants were young women with a history of anaemia and stunting; were 15–19 years at the time of the interview, as shown in **Table 1**.

**Table 1.** Participants’ Characteristic at the Time of Focus Group Discussion.

| Participants' characteristic | n (%) |
| --- | --- |
| **Age** |  |
| 15 | 4 (20.0) |
| 16 | 4 (20.0) |
| 17 | 5 (25.0) |
| 18 | 4 (20.0) |
| 19 | 3 (15.0) |
| **BMI<sup>a</sup>** |  |
| Thinness | 0 (0.0) |
| Underweight | 18 (90.0) |
| Normal | 2 (10.0) |
| Overweight | 0 (0.0) |
| Obese | 0 (0.0) |
| **Anaemic status** |  |
| Mild | 16 (80.0) |
| Moderate | 4 (20.0) |
| Severe | 0 (0.0) |
| **Height-for-Age Z-Score** |  |
| Not stunted | 0 (0.0) |
| Stunted | 20 (100) |
| Severely stunted | 0 (0.0) |
| **Monthly household income** |  |
| Below minimum wage (< USD 128) | 5 (25.0) |
| At minimum wage (USD 128) | 15 (75.0) |
| **Educational status** |  |
| Quit school early | 4 (20.0) |
| At school | 12 (60.0) |
| Graduated | 4 (20.0) |

<sup>a</sup>BMI = Body Mass Index

The young women, anemic and stunted, were arise from a low-income family and have no choice in their life course. They were pressured to quit school early and assist their parents to work in order to sustain their nuclear family’s needs. For example, P10, P11, and P14 stated:

> *Because of a lack of nutrition, the majority of us here are anemic. As others have stated, we realized that the food we eat on a daily basis does not provide balanced nutrition. When compared to protein sources such as meat, fish, and beef, it is more often composed entirely of carbohydrates. We come from a low-income family. The important is, we can eat cheaply but filling. – P10*
>
> *My parents could not afford my school fees, so I dropped out in senior high school. But I do not want to get married too soon; I only started working last week. Even though I received the minimum wage, I believe it was adequate; the important thing is that I can support myself. Now I can buy food with my own money, as opposed to before when I had no choice to only eat twice meals a day. I only ate in the morning and evening, and during the day, I ate dumplings to satisfy a hungry stomach. – P11*
>
> *Like P11, I was unable to pursue my studies after leaving junior high school due to financial constraints by my parents. Since healthy food is prohibitively expensive for us, I eat only two meals per day, with only a tiny amount of food prepared in the home. I have no other choice, already able to eat it; it should be grateful. Ummm, and I began working around a month ago and received the minimum paid – P14*

This study found that young women powerlessness prevailed as the overarching theme in understanding the conditions of anaemia and stunting in the rural setting (see Table 2). Young women in this rural setting are powerless, suffering from financial difficulties and food insecurity, resulting in inadequate nutrition. If parents cannot afford to pay for their children’s education, they believe that early marriage is a viable option for their own and their children’s well-being. Because, by marrying off their children, the parents’ obligations are transferred to their child’s life partner. The powerlessness that young women feel in causing anaemia and stunting children is also expressed in the following P8, P9, P10, P18 and P20 story:

**Table 2.** Summary of Finding.

| Main Theme | Codes |
| --- | --- |
| Young women powerlessness | Financial hardship; household food insecurity; early marriage; nonnutritive food consumption; quit school early; began working for minimum wage; no choice; there are no viable options; monthly household income below the minimum wage; some of the participants had monthly household income at the minimum wage |

> *It is usual in this area for young women from low-income families, when they graduate from high school, they will be prepared for marriage. One of the reasons is to improve people’s lives and eliminate food insecurity. – P8*
>
> *I understand that the food we eat lacks nutritional value, resulting in anaemia. But we can’t do anything because our parents have told us to marry after graduating high school and then live our own lives, hoping that the next generation will not encounter food insecurity like us. – P9*
>
> *My parents persuaded me to marry young to a man of their choice who was thought to help our financial situation. Nonetheless, I was unwilling due to someone’s heartbreaking experience of being forced into early marriage. She was malnourished and anemic during her pregnancy and bleeding during childbirth. My family explained that it occurred because she was not prepared to become pregnant and did not eat healthy foods during her pregnancy due to a lack of money. A public health centre nutritionist has now determined that his child is stunted. Hence, I began working despite the pay being below average. – P18*
>
> *In line with what P18 discussed, my parents felt the same way, planning to marry me with the men right away to alleviate the burden on my parents. But I would not like to marry too soon, so I try to work even though my salary is relatively underpaid and I have not been able to help my parents support my younger siblings. Nevertheless, it appears that I must marry next year; there is no other option. I have been diagnosed with malnutrition and anaemia several times because I do not eat nutritiously and can only eat what is available in the kitchen. – P20*

Household food insecurity causes young women to feel powerless, suffering from anaemia and stunted growth. They can only wish to escape these deplorable conditions to build the best future generation.

> *I hope to continue my education until I graduate from high school. My family’s discussed the possibility of me marrying before graduating from high school because they couldn’t make my life better. According to my understanding, even if I continue to suffer from anaemia, I may experience malnutrition during pregnancy. I absolutely blame myself if my children suffer from stunted growth. – P1*
>
> *My destiny is the same as P1, but I hope that one day I will be able to change it. I’m not even sure I’ll not be malnourished when I get early married. I’m even concerned that I’ll become increasingly anemic and malnourished. I don’t want to have children who are stunted in their development. – P2*
>
> *I can only hope for a brighter future and to bring up a healthy generation. – P3*
>
> *Since years of malnutrition, it’s been not easy to improve my posture, but I believe I can improve my thinking and destiny in the future. – P4*

## Discussion

This study was the first to document another perspective on anaemia and childhood stunting from young Indonesian women in a rural setting. According to our findings, young women stated that their powerlessness were the cause of their anaemia and stunting conditions. Their perception of anaemia and stunting is inextricably linked to their sense of powerlessness thus far. They are powerless in determining a daily diet based on balanced nutrition that is high in iron and other macro-micro nutrients, also the inability to make life decisions hence they quit school early and are forced to married early. The majority of them were from low household income. Furthermore, they are expected to work immediately even if their income is less than the minimum wage and quit school early. In various cross-sectional studies across Indonesia, unsurprisingly insufficient purchasing power and other indices of household wealth are highly connected with child stunting, which is consistent with other research investigations. These elements include poor quality food, inadequate feeding practices, and food and water safety (25–27). The sub-element of poor quality food including poor micronutrient quality, low dietary diversity and low intake of animal source foods, antinutrient content, and lack of energy content of complementary food. Furthermore, the sub-element of food and water safety are contaminated food and water, poor hygiene practices, and unsafe food storage and preparation. A study of the rural population of Brazil found that for 39.7% of people, poverty was an actual fact. A positive relationship was discovered between hemoglobin levels and per capita income as well as a negative relationship between EBIA scores and cardiovascular risk. Individuals experiencing food insecurity, the elderly, and those who do not own a home were found to be more likely to be anemic in a multivariate analysis. Farmers with a per capita income greater than half the minimum wage were less likely to suffer from anaemia. Anaemia was more prevalent in the study group than in previous studies. The disease is associated with risk factors for food insecurity. Improving the determinants of insecurity can aid in the fight against anaemia in children and reproductive women (28).

Young women in our study stated that the daily food consumed was not in compliance with a balanced nutritional menu, excess of carbohydrates and lack of iron also other micronutrients that are required during adolescence to sustain reproductive health. A study in rural Pakistan found that people in rural areas confront multiple barriers to implementing healthy food practices high in nutrients and iron. They also point out that for many years, people have preferred to sell milk and eggs rather than consume them. They spend their earnings on unhealthy food (29). Another study in West Sumatra, Indonesia, found that poverty, food insecurity, dietary restrictions, and a lack of nutritional information were the most serious barriers for the women and families who participated in the investigation (30). Anaemia in Indonesia is primarily caused by a lack of micronutrients such as iron. Despite significant efforts to address anaemia in the community, the problem persists. Health is not solely the responsibility of the government; community participation should be viewed as an effective alternative (31).

Body mass index measurement indicates the condition of a chronic energy deficit (CED) (32). Participants’ low BMI showed that they had a severe energy deficit compared with their peers and had an increased risk of illness and death due to chronic illness and infection since their bodies did not have enough energy to sustain normal growth and functioning. They are more likely to suffer from chronic diseases like diabetes, hypertension, and cancer because of being underweight (33,34). In addition, they are also susceptible to infections. Severely malnourished children or adolescents may develop stunted growth or suffer irreversible cognitive impairments. Therefore, they are unable to develop physical skills efficiently, resulting in poor school performance and lower life prospects (35,36).

A previous study found that food-insecure children aged 15-19 years were nearly three times (13) more likely to have iron deficiency anaemia than food-insecure children, and they may have a more diverse and severe caseload (37). Food insecurity causes anaemia in young and adult women through inadequate consumption of iron, insufficient consumption of micronutrients, thereby facilitating the bioavailability of iron. Early adolescence is a crucial period for iron nutrition since higher iron requirements and the onset of menstrual period (38). Young and adult women require twice as much Fe intake as men because they experience menstrual cycles and pregnancy, which can make them vulnerable to a higher risk of anaemia during food insecurity (8,13,39,40).

Iron intake is stored in the muscles and the spinal cord. If there is insufficient iron, the iron stores in the spinal cord that are used to produce hemoglobin decrease. Hemoglobin transports oxygen from the lungs throughout the body. When Hemoglobin levels fall, free erythrocyte protoporphyrin levels rise, resulting in decreased heme synthesis and erythrocyte size (microcytic erythrocytes). Such circumstances will result in iron deficiency (41). Aside from causing iron deficiency anaemia, iron deficiency can weaken the body’s immune system, allowing infectious diseases to enter the body more easily. Children’s linear growth will be impacted by chronic iron anaemia and infectious diseases.

Therefore, maternal care and assistance for children and adolescents can be viewed as a significant element in preventing anaemia in food insecure households (8). The majority of the participants in this study did not receive food welfare care or support. Other young women’s viewpoints on anaemia and stunting arose as a result of their experiences. The powerlessness of young women to determine their attitude toward consumption nutritious food, their forced dropout from school because they come from a poor economy, and their forced marriage early so that parental responsibility in taking care of children is completed, creates a food insecurity situation that has an impact on the incidence of anaemia and stunting they experience.

## Study Limitations

From study conceptualization to data analysis, the principal investigator was actively involved in all stages of the research. This is accomplished to ensure that the information gathered in the field is of high quality and rich in content. However, there are several limitations to this study, including the fact that the investigator only included young women and did not include single mothers or pregnant young women/adolescents. The interview analysis was then based on the subjective reports of the participants and thus could not be generalized to other settings. Finally, this study does not take into account the other perspectives on anaemia and stunting held by these key stakeholders in Yogyakarta, Indonesia.

## Conclusion

Participants in this study had a good understanding of anaemia and stunting. However, they had lost their autonomy in making decisions about their lives, leaving adolescent girls powerless to meet their nutritional needs. They have endured financial hardship, food insecurity, and the impacts are insufficient nutrition, lack of iron, and stunting. In particular, this study focuses on young women in the lower middle class who face negative social stigma in terms of food security and nutritional adequacy. They lack social support in planning their future, food insecurity, the termination of their education, and finally landing a low-paying job that results in malnutrition. As a result, they are anemic and stunted growth detected. The existing health system needs to reinforce the support for young women to reduce risks in early life and improve their health across the life course. The intervention could include promoting access to nutritious foods, good hygiene, family planning education, and access to health facilities and services

## Data Availability

The datasets generated and/or analyzed during the current study are available upon reasonable request from the corresponding author because of ethical restrictions concerning the sharing of the study data with others without permission from the participants of this study.

## Acknowledgement

The authors express their gratitude to the participants for generously sharing their valuable information. The authors would also like to thank the Ministry of Education and Culture - Research and Technology, in the scheme of implementing the National Research Priority Flagship Program for Universities., Indonesia for research funding assistance, with contract number: 2883/UN27.22/PT.01.03/2021.

## Authors’ Contributions

**Conceptualization:** Giyawati Yulilania Okinarum

**Data curation:** Hardiningsih, Fresthy Astrika Yunita

**Formal analysis:** Hardiningsih, Fresthy Astrika Yunita

**Funding acquisition:** Hardiningsih, Fresthy Astrika Yunita

**Investigation:** Hardiningsih, Afroh Fauziah

**Methodology:** Giyawati Yulilania Okinarum

**Project administration:** Hardiningsih, Fresthy Astrika Yunita

**Resources:** Afroh Fauziah

**Software:** Giyawati Yulilania Okinarum

**Validation:** Giyawati Yulilania Okinarum, Muhammad Hardhantyo

**Writing – original draft:** Giyawati Yulilania Okinarum

**Writing – review & editing:** Giyawati Yulilania Okinarum, Muhammad Hardhantyo

