## Supplemental file for "Understanding anaemia and stunting among young women in a rural setting of Indonesia"

**<u>Supplement 1. Informed consent sheet</u>**

### Explanation Sheet

Hello, we are (Hardiningsih, Giyawati Yulilania Okinarum, Afroh Fauziah, and Fresthy Astrika Yunita) will conduct a research entitled "Remaja Bergerak: An Early Discovery Strategy and Stunting Prevention Since 8000 days of life by Adolescents and Preconceptions in Gunungkidul Regency, Indonesia". This research was sponsored by the Ministry of Eduction and Culture, Research and Technology, through the scheme: National Research Priority Flagship, Acceleration of Stunting Reduction,

This research aims to:

1. Investigate the difficulties and perspectives that adolescents face in implementing with anemia and stunting-free conditions.
2. Investigate adolescents' perceptions and experiences regarding the importance of early detection and prevention of stunting since 8000 days of life in Gunungkidul Regency.

The research team invited you to participate in this research. This research requires about 20 panelists, with a period of 1 to 1.5 hours and will be conducted 1 in-depth interview or 1 Focus GroupDiscussion (FGD).

A. Volunteering to participate in research

You are free to choose to participate in this research without coercion. If you have decided to participate, you are also free to resign/change your mind at any time without being subject to any fines or sanctions.

B. Research procedure

If you are willing to participate in this research, you are required to sign this double consentsheet, one for you to keep, and one for the researcher. The next procedure is:

1. You will be checked for haemoglobin levels using a haemoglobinometer by taking a drop of blood from your fingertrip. Following that, your height and weight will be measured. Midwives will perform examinations and measurements.
2. You will be interviewed by the research team to ask questions that have been in the research instrument.
3. The interview will be conducted for 1 to 1.5 hours.
4. Interviews are conducted once.
5. The researcher will record during the in-dept interview and/or FGD process.
6. The researcher will make a transcript of the interview.

C. Obligations of research subjects

As a research subject, you are obliged to follow the rules or research instructions as written above. If there is anything unclear, you can ask the researcher further.

D. Risks and Side Effects

The risk for the subject could be data leakage. To prevent data leakage and maintain the confidentiality of the subject's identity, each subject will be given a data source code and in the publication of research results will be published without the identity of the research subject.

E. Benefits

You will benefit from this research in the following way: Skill development by utilizing local food ingredients with the potential to reach the nutrition needs, and entrepreneurship development by marketing and selling regionally superior products, can improve the economic and social status of the Gunungkidul Regency area, particularly in adolescents who already have stunting or anemia as a result of food insecurity.

F. Confidentiality

All information relating to the identity of the research subject will be kept confidential and will only be known by researcher and the research staff. The results of the research will be published without the identity of the research subjects.

G. Compensation

Participants will receive compensation two hundred thousand rupiah.

H. Financing

All research-related costs will be borne by researchers and sponsors.

I. Additional Information

You can ask all the things that are not clear about this research. If you need an explanation, you can contact Giyawati Yulilania Okinarum, S.ST., M.Keb. at (phone number) at the Department of Professional Midwives Program, Faculty of Science, Universitas Respati Yogyakarta, Jl. Raya Tajem KM 1,5 Sleman Yogyakarta, Indonesia.

You can also ask about research to the Medical and Health Research Ethics Committee of Faculty of Healths Science Universitas Respati Yogyakarta (Phone. 0274-4437888 or +62822-1023-7139.

**Statement of Informed**
**Consent**

I, voluntarily participated in this research launched by the Universitas Respati Yogyakarta and Universitas Sebelas Maret.

I have read or received an explanation, am fully aware of and understand the objectives, benefits, and risks that may arise in the research, have been given the opportunity to ask questions and have satisfactorily answered ones, and may withdraw from participation at any time. Then I agree to take part in this study, titled:

*"Remaja Bergerak: An Early Detection Strategy and Prevention of Stunting by Adolescents and Preconceptions in Gunungkidul Regency Since 8000 Days of Life"*

I participated in this research voluntarily, without any pressure or coercion from anyone. I will be given a copy of the explanation sheet and consent form I signed for my file.

By signing this form, I agreed to participate in this research.

Subject signature: Date __ / __ / ____

Clear Name:

Witness signature:

Full Name :

**General Information (Filled out by interviewer)**

1. Interviewer :
2. Minutes :
3. Date :
4. Time : Start___________and Finish _______
5. Location :
6. Use

Voice Recorder : (Yes/No), If Not, the reason:

**Characteristics of Informants (Filled by interviewer)**

7. Name : ___________________
8. Age : ______________
9. Gender : ____________________
10. Education : _______________________
11. Household income : ___________________
12. Haemoglobin level : ___________________
13. Weight and height : ______________
14. Body mass index : ______________
15. Informant code : ________

**Supplement 2. Interview Guidelines**

**Procedures for opening an interview**

Good morning/ noon/ afternoon, let me introduce us, I am … and my partner… . Thank you for taking the time. We will ask some questions, and please answered freely expressing the opinion. There are no right or wrong answers. This interview will last from 1 hour to 1.5 hour. To facilitate transcription, this interview will be recorded. We will keep all the information; no names and titles will be released. Do you agree to have this interview recorded?

Do you have any questions before we start?

| Issue(s) explored | Question | Probing/ Dunning |
|---|---|---|
| Description about stunting and anemia | Can you tell us about stunting and anemia? | Does anemia affect stunting in future?<br><br>How did you feel when you were diagnosed with stunting and anemia? |
| Early detection and prevention in stunting | Tell us about the importance of early detection, prevention in stunting, and about the early marriage | How important is early detection and prevention of stunting since 8000 days of life?<br><br>Do you take part in early detection and early detection of stunting and anemia in your community? |
| Implementation challenges | Tell us about the challenges you've faced in reaching your nutritional needs so far! | How hard have you worked to fulfill your nutritional needs? |
| Implementation strategies | What strategies are you taking to get out of this food insecurity? | How effective have your strategies been? |
